# Dynamics of neutralizing antibody responses in acute-phase COVID-19: A potential relationship between disease progression and rapid neutralizing antibody response

**DOI:** 10.1101/2021.02.06.21251246

**Authors:** Hitoshi Kawasuji, Yoshitomo Morinaga, Hideki Tani, Miyuki Kimura, Hiroshi Yamada, Yoshihiro Yoshida, Yusuke Takegoshi, Makito Kaneda, Yushi Murai, Kou Kimoto, Akitoshi Ueno, Yuki Miyajima, Koyomi Kawago, Yasutaka Fukui, Ippei Sakamaki, Yoshihiro Yamamoto

**Author notes:** **Correspondence:** Yoshihiro Yamamoto, MD, PhD, Department of Clinical Infectious Diseases, Graduate School of Medicine and Pharmaceutical Sciences, University of Toyama, 2630 Sugitani, Toyama, 930-0194, Japan. ***ICMJE Statement*** All authors met the ICMJE authorship criteria.

## Abstract

**Introduction:** Adaptive immunity to severe acute respiratory syndrome coronavirus 2 (SARS-CoV-2) dynamics remain largely unknown. The neutralizing antibody (NAb) levels in patients with coronavirus disease 2019 (COVID-19) are helpful for understanding the pathology.

**Patients and Methods:** Using SARS-CoV-2 pseudotyped virus, serum sample neutralization values in symptomatic COVID-19 patients were measured using the chemiluminescence reduction neutralization test (CRNT). At least two sequential serum samples collected during hospitalization were analyzed to assess NAbs neutralizing activity dynamics at different time points.

**Results:** Of the 11 patients, four (36.4%), six (54.5%), and one (9.1%) had moderate, severe, and critical disease, respectively. Fifty percent neutralization (N50%-CRNT) was observed upon admission in 90.9% (10/11); all patients acquired neutralizing activity 2–12 days after onset. In patients with moderate disease, neutralization was observed at earliest within two days after symptom onset. In patients with severe-to-critical disease, neutralization activity increased, plateauing 9–16 days after onset. Neutralization activity on admission was significantly higher in patients with moderate disease than in patients with severe-to-critical disease (relative % of infectivity, 6.4% vs. 41.1%; *P*=.0011).

**Conclusions:** Neutralization activity on admission inversely correlated with disease severity. The rapid NAb response may play a crucial role in preventing the progression of COVID-19.

## Introduction

Understanding the dynamics and key features of adaptive immunity to severe acute respiratory syndrome coronavirus 2 (SARS-CoV-2) is essential for controlling the coronavirus disease 2019 (COVID-19) pandemic; however, the details of the emergence, establishment, and long-lasting memory of adaptive immunity in response to SARS-CoV-2 remain largely unknown. Antibodies are important elements of the adaptive immune response. Recent evidence shows that recovered COVID-19 patients can generate antibodies, albeit at variable levels, that specifically bind to various structural proteins of SARS-CoV-2 particles shortly after the onset of disease^[1-3]^. Among these virus-specific antibodies, only those capable of blocking SARS-CoV-2 spike (S) protein-mediated viral attachment and/or entry of host cells, called neutralizing antibodies (NAbs), play an important role in virus clearance and have been considered as key immune products in the protection against or treatment of viral diseases. Severe COVID-19 can be associated with high neutralization titers in the sub-acute or later phases^[4,5]^; however, whether the acquisition of neutralizing antibodies in the acute phase is related to disease severity is unclear.

The level of NAbs has also been used as a gold standard for evaluating the efficacy of vaccines. However, the measurement of NAb levels is needed to deal with infectious virus and active cells, which makes it a difficult procedure with respect to the problems of standardization and the need for specialized laboratory equipment with BSL-3 facilities.

To circumvent the need for BSL-3 laboratories, we recently developed a fundamental NAbs detection method for SARS-CoV-2 based on the chemiluminescence reduction neutralization test (CRNT) and VSV-based pseudotyped viruses with SARS-CoV-2 S (Sfullpv) or truncated S proteins (St19pv) to evaluate the expression and neutralization values of NAbs against SARS-CoV-2^[6]^.

In this study, at least two sequential serum samples collected on admission and during hospitalization from confirmed symptomatic COVID-19 patients with different disease severities were analyzed to assess the dynamics of the neutralizing activities of NAbs at different time points and determine the clinical significance of the measurement results using our fundamental NAbs detection method.

## Material and Methods

### Study design

We performed a single-center study at the Toyama University Hospital. Hospitalized adult symptomatic COVID-19 patients with different disease severities, in which at least two sequential serum samples were collected during hospitalization, were included in this study. The patients were tested for SARS-CoV-2 RNA in the serum and nasopharyngeal (NP) swabs collected on or around admission. Given that viral culture was not performed to determine the viability of the virus, the presence of viral RNA in the serum is referred to as RNAemia instead of viremia^[7]^. To study the dynamics of the neutralizing antibody response, blood samples from symptomatic patients were collected on admission, and subsequently when clinically needed during hospitalization.

### Data Collection

A retrospective chart review was performed for all individuals in the study to identify basic demographic data, medical history, clinical presentation, and hospital admission. Severity was divided into four categories: mild (symptomatic patients without pneumonia), moderate (pneumonia patients without required oxygen supplementation), severe (requiring oxygen supplementation), and critical (requiring invasive mechanical ventilation, presence of shock and/or multiple organ dysfunction).

### Neutralization Assay

Neutralization of the human serum against pseudotyped viruses was measured based on a previous report^[6]^. Briefly, Vero cells were treated with two hundred-fold dilutions of sera from in-hospital patients with COVID-19 and then inoculated with pseudotyped VSVs bearing the S protein, the 19-amino acid-truncated S protein of SARS-CoV-2 or VSV-G. The infectivity of the pseudotyped viruses was determined by measuring luciferase activity after 24 h of incubation at 37°C, based on our preliminary study^[6]^, and the number of days after admission or onset until the first result of 50% neutralization in the CRNT (N50%-CRNT) was recorded.

### Statistical analysis

Continuous and categorical variables are presented as median (interquartile range [IQR]) and n (%), respectively. We used the Mann–Whitney U test, χ^2^ test, or Fisher’s exact test to compare the differences between the patients with moderate and severe/critical disease. Data were analyzed using JMP Pro version 15.0.0 software (SAS Institute Inc., Cary, NC, USA). The time courses of the neutralization activities were also analyzed using nonlinear regression in JMP Pro version 15.0.0 software (SAS Institute Inc., Cary, NC, USA).

### Ethical approval

This study was performed in accordance with the Helsinki Declaration and was approved by the ethical review board of the University of Toyama (approval No.: R2019167). Written informed consent was obtained from all patients.

## Results

Of the 11 in-hospital symptomatic patients enrolled in this study, ten patients recovered and were discharged from the hospital. The median age of patients was 78 years (range, 59–100 years); three (27.3%) patients were male, and eight (72.7%) patients were female. Five (45.5%), two (18.2%), and one (9.1%) patients had a history of hypertension, cardiovascular disease, and diabetes, respectively. Four patients (36.4%) had moderate disease, six (54.5%) had severe disease, and the remaining patients (9.1%) had critical disease. For the neutralization test of the first sample, N50%-CRNT was observed on admission in 90.9% (10/11) of all patients and thereafter observed in all patients from 6 and 12 days after admission and onset, respectively. The median time from onset to first achievement of N50%-CRNT was 8 days (range, 2–12 days).

Based on the sequential analysis during hospitalization, a total of 79 serum samples from 11 patients were assessed by measuring the neutralization activities. The median number of CRNTs performed in each patient was seven (range, 2–22), and the number of days between the CRNT and onset ranged from 2 to 33 days (Figure 1 and Supplemental Figure 1). The median length of hospital stay was 46 days (range, 14–58 days) (Table 1). Regarding the evaluation of the kinetics of neutralization activities, there were different characteristics according to disease severity. In patients with moderate disease (Figure 1), the degree of neutralization activity in the first samples collected on admission was high, and neutralization was observed two days after symptom onset in patient #11 (Supplemental Figure 1). In contrast, in severely critically ill patients, the degree of neutralization activity in the first samples collected on admission was relatively low and gradually increased to reach a plateau approximately 9–16 days after onset. Notably, when comparing neutralization activities on admission between patients with moderate and severe-to-critical disease, the degree of neutralization activity of St19pv in the moderate patients was significantly higher than that in the patients with severe-to-critical disease (6.4 ± 3.4% vs. 41.1 ± 24.6%; *P*=.0011) (Table 1).

**Table 1.**
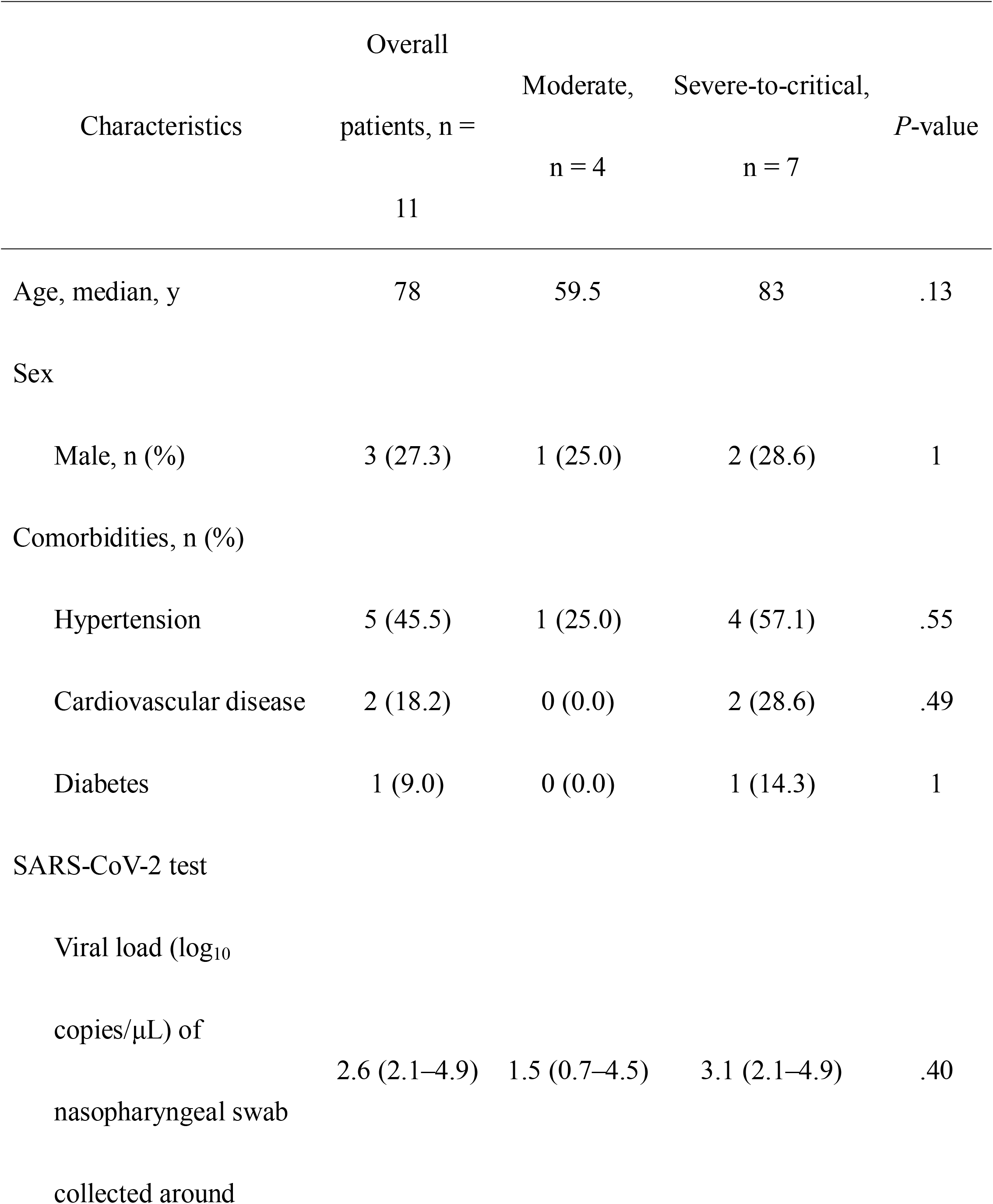
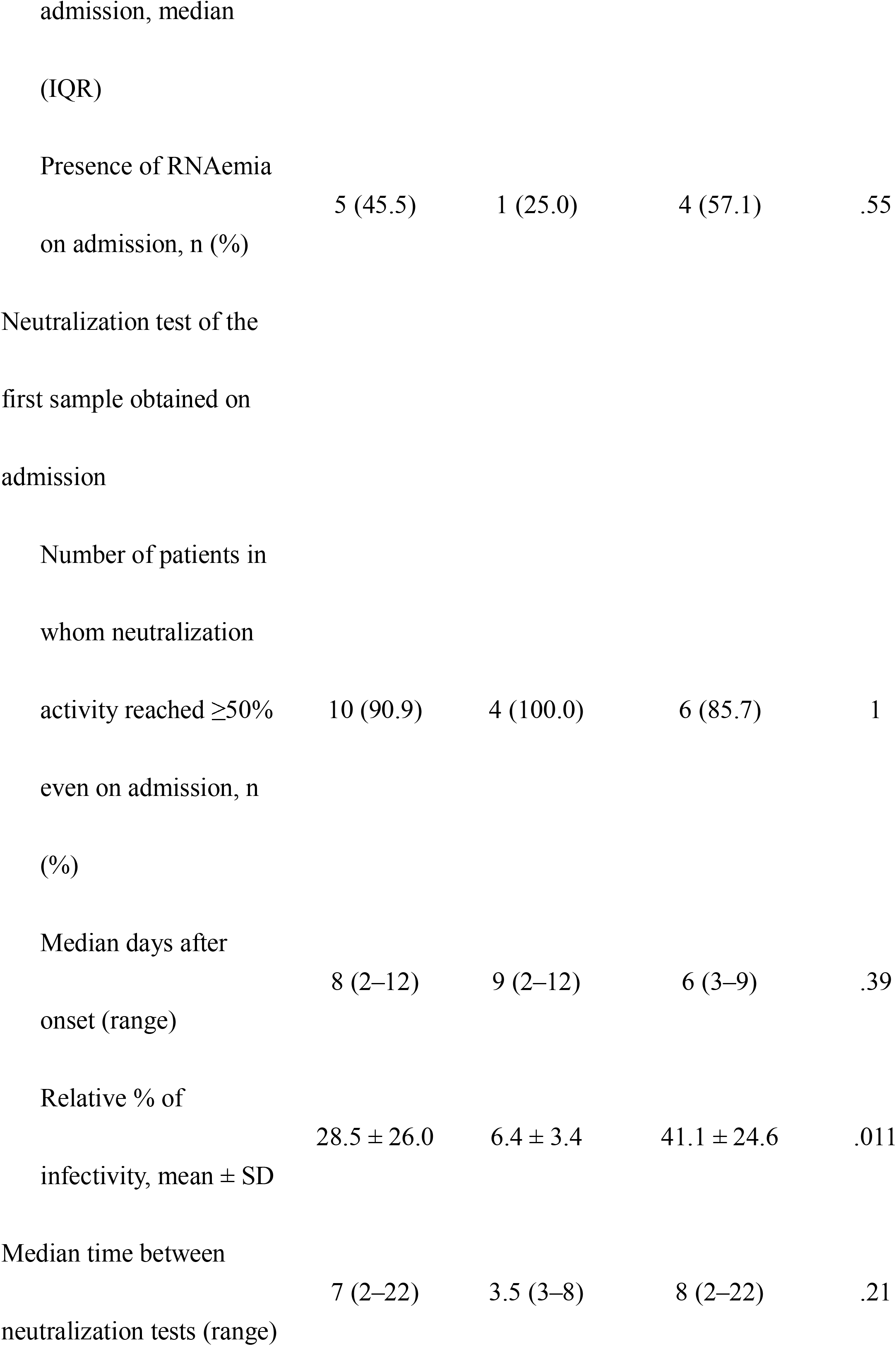
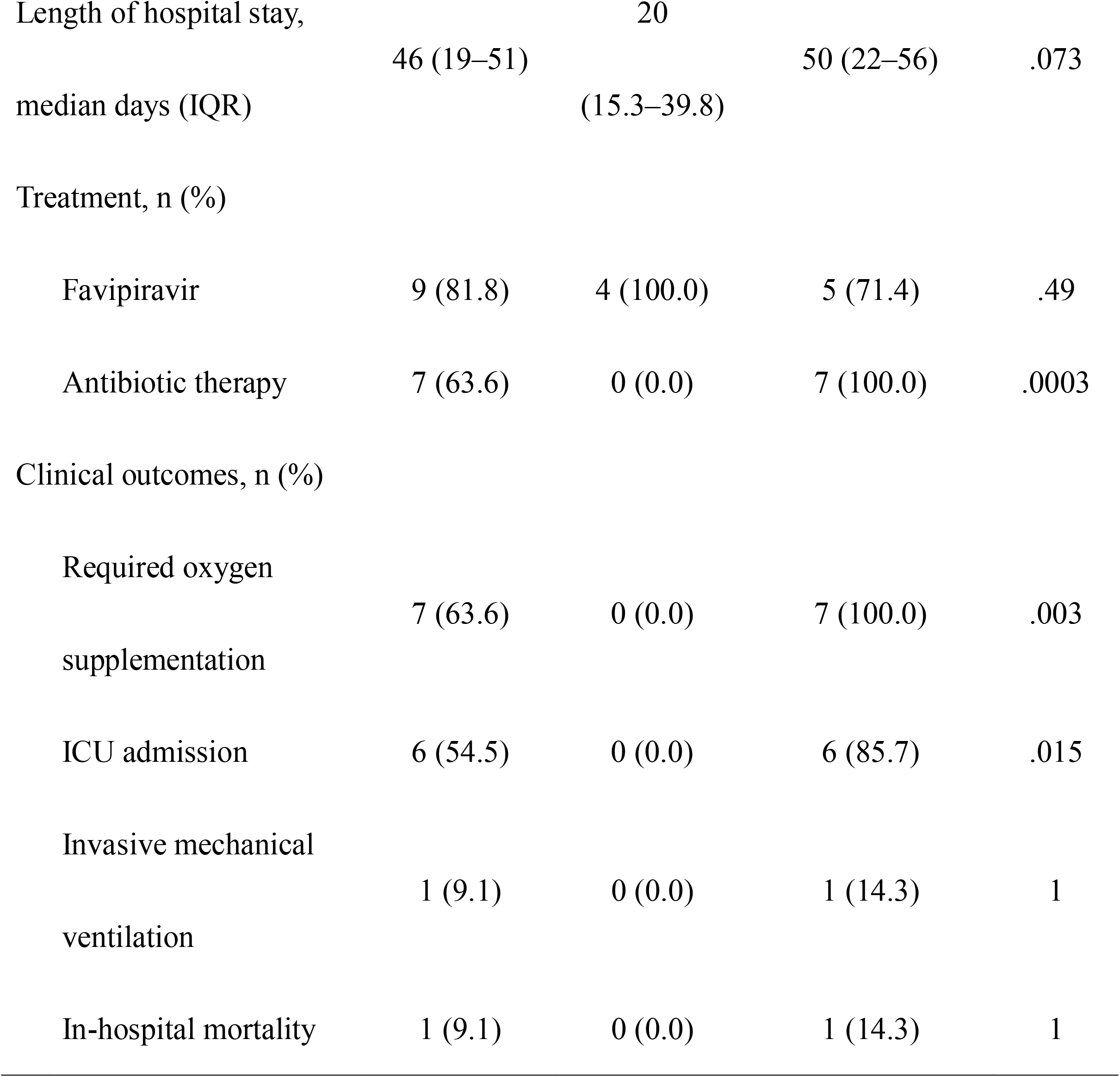
Demographic, clinical characteristics, results of SARS-CoV-2 test, and neutralization antibody activities on admission of eleven patients stratified by severity

**Figure 1.**
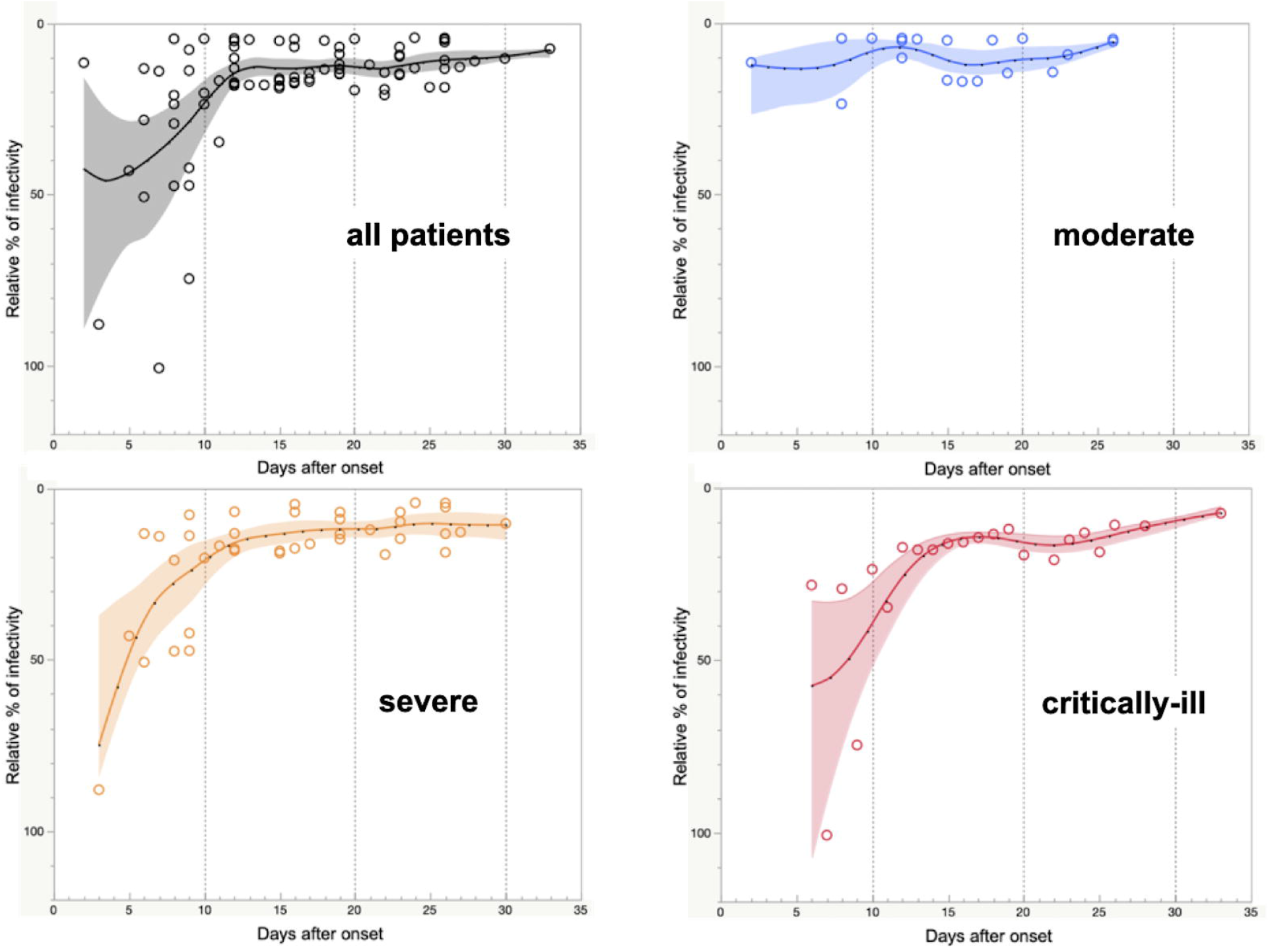
Trends in neutralization activities in symptomatic patients stratified by severity. Nonlinear regression models of all (black), moderate (blue), severe (orange), and critically ill patients (red). The models were calculated using the mean data of the patients stratified by severity. Solid curves are spline curves fitted to the mean with a λ of 0.05. Shading indicates the 95% confidence interval of each model.

## Discussion

It is still unknown when sufficient immunity against SARS-CoV-2 is obtained during the course of an infection. In the present study, we confirmed that patients who recovered from COVID-19 acquired neutralizing activity 2–12 days after onset based on CRNT. However, it is not known which type of antibody predominantly plays a role in neutralization because the CRNT result reflects the comprehensive action of a variety of antibodies. In a recent study on COVID-19, Sterlin et al.^[8]^ suggested that neutralizing capacity rapidly increased and plateaued by day 10 post-symptom onset, which is consistent with our results for patients with moderate and severe disease. They also indicated the possibility that early SARS-CoV-2-specific humoral responses were dominated by IgA. Another study reported that seroconversion occurred 2 days after symptom onset^[9]^, supporting the early acquisition of neutralizing activity in patient #11 (Supplemental Figure 1).

To date, the relationship between disease severity and the magnitude of the SARS-CoV-2-specific NAbs responses in patients with COVID-19 is largely unknown. A recent study demonstrated a positive correlation between the magnitude of NAbs responses and disease severity; in particular, the highest NAbs capacity in the sera was found in patients with severe disease who recovered from COVID-19^[1]^. These results were not consistent with our results, but this discrepancy may be attributed to the different phases, the acute and convalescent phases, in which the NAbs response was measured.

In the acute phase on admission, our study found that the degree of neutralization activity in patients with moderate disease was significantly higher than that in patients with severe-to-critical disease. It has been reported that NAbs appear earlier at higher levels in patients with severe or moderate disease than in those with mild disease^[10]^; however, our findings clearly indicate the difference in the induction of the neutralizing response between moderate and severe-to-critical disease. These results suggest that the acquisition of NAbs in the early stages of infection may be related to disease severity. Because severity positively correlates with SARS-CoV-2 RNAemia^[7]^, the acquisition of NAbs, including mucosal immunity, can modify early COVID-19 pathology. In contrast, in the convalescent phase in COVID-19 patients, high NAbs capacity in elderly patients and/or those with severe disease as previously reported^[1,11]^, which is mainly due to specific IgG, could be a result of the severe inflammatory response. Therefore, the specific background related to the ability of NAbs should be verified.

This study had several limitations. First, the number of involved patients was very small and selected using consecutive instead of random sampling. Thus, representativeness is relatively insufficient, and the samples could only represent the general situation to a certain extent. Second, the subjects mainly had moderate and severe disease, and only one critically ill patient was included. The NAb response in asymptomatic infections and mild patients requires further exploration. Third, the meaning of the quantitative values of CRNT results remains controversial. To make the value clinically meaningful, continuous improvement of our method and comparison with clinical findings are required.

## Conclusions

In conclusion, this study showed that neutralizing antibody activities were observed in all patients with symptomatic COVID-19, even in the early stage of illness, and the degree of neutralization activity on admission was significantly higher in patients with moderate disease than in those with severe-to-critical disease. The present study highlights that a rapid NAb response may play a crucial role in preventing severe progression of COVID-19.

## Data Availability

All relevant data are within the manuscript.

## Author contributions

HK, YMo, HT, and YY designed the study. HK, YMo, HT, MKi, HY, and YYo performed the experiments. HK, YT, MKa, YMu, KKo, AU, YMi, KKa YF, and IS collected the samples and contributed to the completion of the study. HK and YMo contributed to data analysis and wrote and edited the manuscript. All the authors approved the final version of the manuscript.

---

**Supplemental Figure 1.**
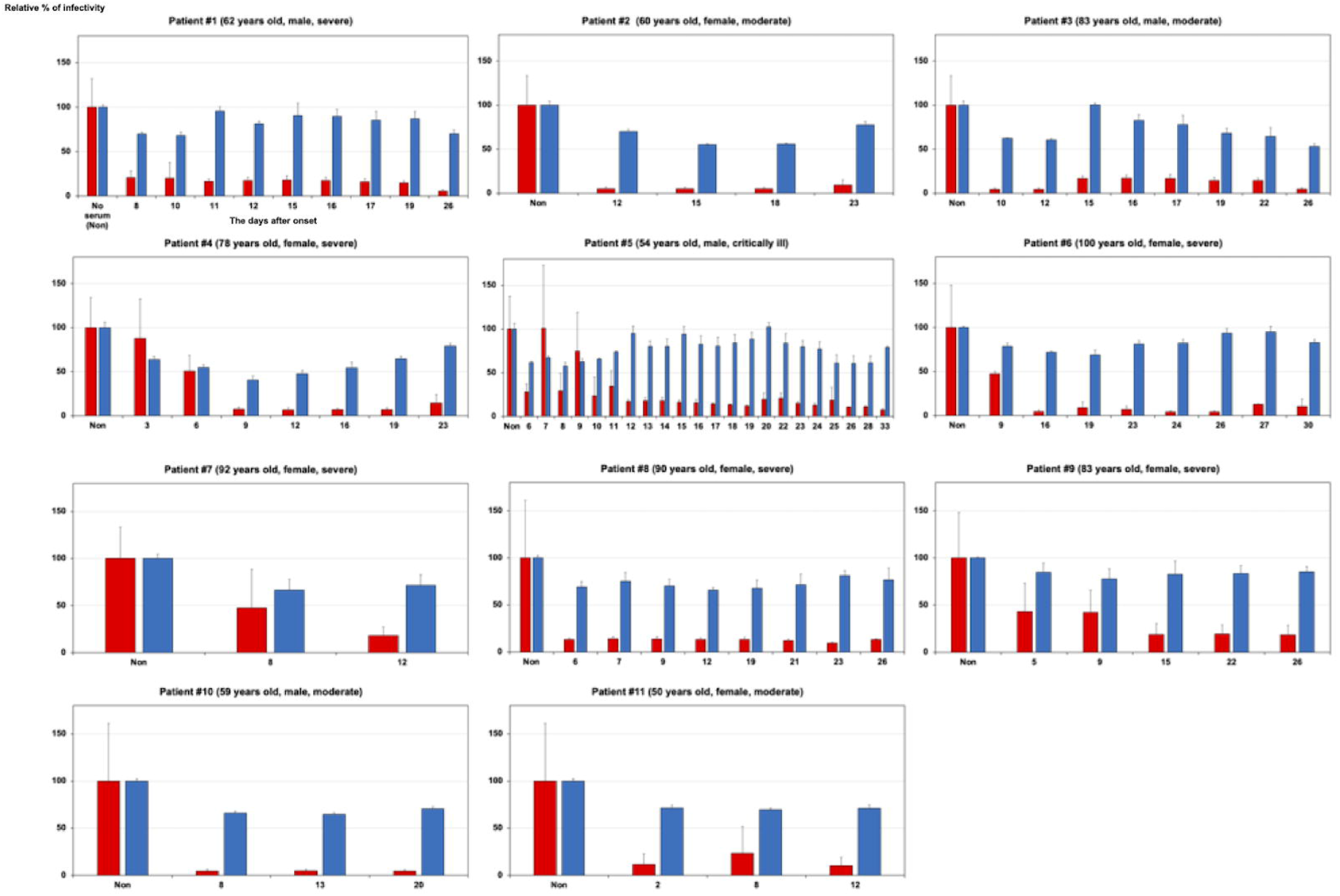
Neutralization of St19pv and VSVpv infections through sera from eleven hospitalized patients #1–#11 with COVID-19. The pseudotyped viruses were preincubated with two a hundred-fold dilution of sera from eleven (#1–#11) hospitalized COVID-19 patients. Thereafter, Vero cells were infected with pseudotyped viruses. Infectivities of pseudotyped viruses were determined by measuring luciferase activities 24 h post-infection. The results are from three independent assays with error bars representing standard deviations.

## References

1. Chen, X. et al. Disease severity dictates SARS-CoV-2-specific neutralizing antibody responses in COVID-19. Signal Transduct. Target. Ther. 5(1), 180 (2020). 10.1038/s41392-020-00301-9, https://www.ncbi.nlm.nih.gov/pubmed/32879307

2. Long, Q. X. et al. Antibody responses to SARS-CoV-2 in patients with COVID-19. Nat. Med. 26(6), 845–848 (2020). 10.1038/s41591-020-0897-1, https://www.ncbi.nlm.nih.gov/pubmed/32350462

3. Amanat, F. et al. A serological assay to detect SARS-CoV-2 seroconversion in humans. Nat. Med. 26(7), 1033–1036 (2020). 10.1038/s41591-020-0913-5, https://www.ncbi.nlm.nih.gov/pubmed/32398876

4. Garcia-Beltran, W. F. et al. COVID-19-neutralizing antibodies predict disease severity and survival. Cell 184(2), 476–488.e11 (2021). 10.1016/j.cell.2020.12.015, https://www.ncbi.nlm.nih.gov/pubmed/33412089

5. Legros, V. et al. A longitudinal study of SARS-CoV-2-infected patients reveals a high correlation between neutralizing antibodies and COVID-19 severity. Cell. Mol. Immunol., 1-10 (2021). 10.1038/s41423-020-00588-2, https://www.ncbi.nlm.nih.gov/pubmed/33408342

6. Tani, H. et al. Evaluation of SARS-CoV-2 neutralizing antibodies using a vesicular stomatitis virus possessing SARS-CoV-2 spike protein. Virol. J. 18(1), 16 (2021). 10.1186/s12985-021-01490-7, https://www.ncbi.nlm.nih.gov/pubmed/33435994

7. Kawasuji, H. et al. SARS-CoV-2 RNAemia with higher nasopharyngeal viral load is strongly associated with severity and mortality in patients with COVID-19. medRxiv, 2020.2012.2017.20248388. 10.1101/2020.12.17.20248388 (2020)

8. Sterlin, D. et al. IgA dominates the early neutralizing antibody response to SARS-CoV-2. Sci. Transl. Med. 13(577) (2021). 10.1126/scitranslmed.abd2223, https://www.ncbi.nlm.nih.gov/pubmed/33288662

9. Yu, H. Q. et al. Distinct features of SARS-CoV-2-specific IgA response in COVID-19 patients. Eur. Respir. J. 56(2) (2020). 10.1183/13993003.01526-2020, http://www.ncbi.nlm.nih.gov/pubmed/32398307

10. Jeewandara, C. et al. SARS-CoV-2 neutralizing antibodies in patients with varying severity of acute COVID-19 illness. Sci. Rep. 11(1), 2062 (2021). 10.1038/s41598-021-81629-2, https://www.ncbi.nlm.nih.gov/pubmed/33479465

11. Wang, X. et al. Neutralizing Antibodies Responses to SARS-CoV-2 in COVID-19 inpatients and convalescent patients. Clin. Infect. Dis. (2020). 10.1093/cid/ciaa721, https://www.ncbi.nlm.nih.gov/pubmed/32497196

